# Continuous at-home wearables and machine learning sensitively track function and progression in pediatric and adult Friedreich’s ataxia

**DOI:** 10.64898/2026.11.23.26368776

**Authors:** Kaitlyn Arrambide, Faye X. Yang, Jennifer Farmer, Myriam Rai, Victoria Profeta, David Lynch, Anoopum S. Gupta

**Author notes:** Correspondence to: Anoopum S. Gupta 101 Merrimac St Suite 300, Boston, MA 02114. equal contributions (Kaitlyn Arrambide, Faye X. Yang).

## Abstract

Friedreich’s ataxia (FRDA) is the most common hereditary ataxia, characterized by progressive cerebellar and sensory ataxia, proprioceptive loss, and upper motor neuron involvement, with onset typically in childhood. Current outcome measures used in clinical trials are limited by rater subjectivity, coarse scoring, infrequent in-person assessments, and inability to assess behavior continuously in naturalistic settings. We tested whether submovement-based measures derived from accelerometers worn continuously at home could sensitively capture motor impairment and disease progression in FRDA across a range of ages and disease severity. 95 participants (57 FRDA, 38 controls; age 2-75) wore GENEActiv wrist and ankle sensors for one week per timepoint; 60 completed two or more timepoints spanning up to 2.2 years. 85 movement features were extracted including submovement kinematics and activity intensity. The FRDA cohort included ambulatory and non-ambulatory participants spanning early, mid, and late stages of disease. Five machine-learned composite models were evaluated, including FRDA-specific models, and models previously trained on other populations. Sensor measures were evaluated for relationships with clinician-rated and patient-reported measures, reliability, and sensitivity to progression. Ankle sensor measures demonstrated strong cross-sectional correlations with Friedreich’s Ataxia-Functional Disability Staging (|r| = 0.66–0.87), Modified Friedreich’s Ataxia Rating Scale total (|r| = 0.53–0.73), Friedreich’s Ataxia-Activities of Daily Living (|r| = 0.58–0.74), and Friedreich’s Ataxia-Health Index total (|r| = 0.43–0.57), with high within-week reliability (ICC = 0.89–0.96) and large differences between FRDA and control groups (|e.s.| = 2.0–2.6). FRDA-specific composite outcomes showed significant longitudinal sensitivity to disease progression (|MSDR| = 0.67–0.89), whereas patient global impression of change did not capture progression. At-home ankle accelerometer data yield reliable, ecologically valid measures reflecting clinician-rated and patient-reported outcomes, and capture disease progression in FRDA. These results support remote wearables as sensitive digital outcome measures for FRDA clinical trials and disease monitoring.

**Research In Context:** *Evidence before this study:* We searched PubMed and Google Scholar for articles published up to September 2026 using the terms “Friedreich’s ataxia”, “wearable”, “accelerometer”, “digital biomarker”, and “motor outcome measure”. Currently, disease severity and progression in FRDA are measured through in-person, clinician-rated exams and functional assessments. However, these scoring systems are coarse, have limited sensitivity across the full disease spectrum, and may not reflect day-to-day variability in motor function in individuals with FRDA. Prior wearables studies in FRDA using metrics such as step count and vector magnitude have demonstrated the ability to capture progression from proximal sensor locations but were limited by behavioral variability and inability to capture fine-grained movement patterns. Other studies using pendant and wrist sensors have shown correlations with FRDA severity markers but did not collect lower limb sensor data or assess longitudinal sensitivity. No prior studies have evaluated at-home ankle-based submovement measures across the full FRDA age and severity spectrum, including pediatric and non-ambulatory individuals.

*Added value of this study:* In a remote wearable study consisting of 57 individuals with FRDA spanning early, mid, and late stages of disease, and 38 controls, we show that submovement-based measures derived from ankle accelerometer data collected continuously at home correlate strongly with clinician-rated and patient-reported outcomes, have high within-week reliability, and demonstrate large differences between FRDA and control groups. FRDA-specific machine-learned composite models showed the strongest sensitivity to longitudinal disease progression and outperformed models trained on data from other neurologic populations. Sensor-based measures captured progression that patient-reported global impressions of change did not, underscoring the sensitivity and objectivity of digital measures in FRDA.

*Implications of all the available evidence:* To our knowledge, this is the first study to generate digital outcome measures across the full spectrum of FRDA using continuously worn, at-home wearable sensors. By demonstrating the reliability, ecological relevance, and sensitivity of our sensor-based and submovement-derived digital measures, this study establishes a framework for remote digital outcome measures in FRDA clinical trials.

## Introduction

The development of new therapies for Friedreich’s ataxia (FRDA), the most common hereditary ataxia, has increased in recent years. Several disease modifying therapies are in or nearing clinical trials,^1^ and omaveloxolone (Skyclarys™) achieved FDA approval in February of 2023 as the first effective therapy for FRDA.^1,2^ These trials underscore the need for sensitive and reliable outcome measures that reflect real-world behavior (i.e., ecologically valid) to accurately and efficiently evaluate new therapeutic modalities. Despite this progress, gaps remain in the tools available to measure progression and determine the effectiveness of treatment in naturalistic settings.

FRDA is an autosomal recessive neurodegenerative disease caused by biallelic GAA repeat expansions within the *FXN* gene, leading to decreased production of the mitochondrial protein frataxin.^3^ Downstream effects of deficient frataxin levels include progressive cerebellar and sensory ataxia, degeneration of corticospinal tracts, cardiomyopathy, and scoliosis. These symptoms progressively affect daily life, causing difficulties with gait and limb coordination and eventually leading to loss of independent ambulation.^4–6^ Capturing early behavioral changes in FRDA is particularly difficult. Symptoms in the initial stages of disease can be subtle and overlap with typical pediatric motor development,^7^ making it challenging to detect meaningful change with existing tools. Thus, sensitive measurement tools that can capture motor function across the full disease spectrum are needed to improve disease assessment in FRDA.

Currently, disease severity and progression in FRDA are measured through in-person, clinician-rated exams and functional assessments. The most widely used clinical outcome measure in FRDA is the Friedreich’s Ataxia Rating Scale (FARS) and its modified version (mFARS),^8^ which assess bulbar, upper limb, lower limb, and upright stability functions (See Supplementary Materials for validation and reliability references). Such clinician-rated scales are prone to rater subjectivity and inter-rater variability, reducing their sensitivity to capture change over time or in response to a therapy.^9,10^ The scoring systems for these scales are inherently coarse, as they are constrained by limits of human perception and may miss small, continuous changes in motor function.^9^ They are prone to floor and ceiling effects, limiting sensitivity at the mildest and most severe stages of disease.^8,11,12^ Additionally, assessments conducted during in-person clinic visits are brief and discrete, capturing only a snapshot of functional abilities rather than day-to-day variability.^1,9,11^ This raises questions about ecological validity of the assessments, as performance on prescribed tasks may not reflect an individual’s functional capacity in everyday life.^9,10^ Furthermore, in children, natural development of motor function can overlap with early neurodegenerative symptoms, and standardized prescribed tasks may yield inconsistent results.^7,10^ Standardized tasks such as the Timed 25-Foot Walk (T25-FW) (See Supplementary Materials) and 9-Hole Peg Test (9-HPT)^13^ may be influenced by motivation, fatigue, and testing conditions and therefore cannot account for behavioral patterns at a broader level.^9,11,12^ Lastly, reliance on in-person, clinician-rated assessments poses logistical difficulties for a geographically diverse patient population, and limits the naturalistic assessment of functional capacity and disease progression.^14,11^

Wearable sensors worn during natural behavior at home offer the potential for objective, precise, and ecologically valid measures of motor function that capture how individuals move in daily life.^14,15^ Compared to discrete tasks administered in the clinic, continuous accelerometer-based sensor monitoring is less affected by motivational and performance variability, allowing for data aggregation that stabilizes such effects. Accelerometer-based monitoring also enhances accessibility through remote data capture, reducing burden on patients and clinical teams.

Moreover, analysis of natural behavior may be applicable regardless of ambulatory status or age, supporting accessible assessment across the spectrum of disease severity in FRDA. One promising analytic framework for extracting motor information from accelerometer data is the characterization of submovements, the building blocks theorized to underlie voluntary movements.^16,17^ In ataxias, cerebellar dysfunction disrupts coordination of submovements, causing them to become smaller, slower, and less regular.^17–20^ Because submovement properties are derived from the intrinsic properties of all movements, they can reflect movement quality independent of the specific activity and capture movement impairment across age and functional status. Prior work has shown that analysis of submovements during natural behavior sensitively captures motor impairment and disease progression across several neurologic populations including spinocerebellar ataxias,^9,21^ ataxia-telangiectasia (A-T),^10^ Parkinson’s disease,^22^ and amyotrophic lateral sclerosis (ALS).^23^

In FRDA, prior wearables research has used metrics such as step count^27^ and vector magnitude^26^ recorded from areas including the waist, wrist, or triceps. These studies demonstrated the ability to capture progression but are limited by several factors. Aggregate metrics like step count are sensitive to variability in sensor wear time, choice of daily activities, motivation, and lifestyle.^24^ Additionally, seasonal effects on activity patterns can add variability unrelated to disease state^24^, and summary activity metrics like step count and vector magnitude may not capture fine-grained movement patterns of the limbs most affected in FRDA.^24,25^ More recently, multi-sensor approaches using pendant and wrist sensors worn during daily activities have shown significant correlations with mFARS and proxies for disease severity in FRDA, including GAA repeat length and frataxin protein levels.^26^ However, this approach did not include lower limb sensor data, lower limb function was not directly evaluated for non-ambulatory individuals, and longitudinal properties were not assessed.

Here we evaluate whether submovement-based measures derived from continuous at-home wearable sensors placed on the ankle and wrist can produce reliable, ecologically valid, and sensitive outcome measures across a wide age and severity spectrum in FRDA. We hypothesize that machine-learned composite models specific to FRDA will predict severity and progression over time and be applicable across the full spectrum of FRDA, including pediatric and non-ambulatory individuals.

## Methods

### Recruitment and Consent

Participants took part in the study between June 2023 and January 2026. The study was approved by the Partners Healthcare Research Committee Institutional Review Board (no.2019P002752). Informed consent was obtained from all participants in accordance with the Declaration of Helsinki. Eligibility criteria required that all participants: (i) have a confirmed diagnosis of FRDA or qualify as a healthy control; (ii) be sufficiently proficient in English; (iii) be at least 2 years of age; and (iv) have access to a computer and internet. Participants over 75 years of age were excluded to reduce heterogeneity related to aging (see Supplementary Figure 1).

Ninety-five individuals were included in the analysis: 57 FRDA (1 pre-symptomatic) and 38 controls (Table 1). Pre-symptomatic was defined as genetically confirmed FRDA without self or caregiver-reported ataxia symptoms during participation. The one pre-symptomatic individual was not analyzed separately. The sample represented broad geographical diversity, with participants residing across 33 states and 3 countries, including the United States, Canada, and the United Kingdom. Individual-level demographic and clinical information is shown in Supplementary Table 1. Sixty of 95 participants (35 FRDA, 25 controls) had longitudinal data across two or more timepoints (Table 1 and Supplementary Fig. 1).

**Table 1.** Participant demographic and clinical information.

|  | Cross Sectional Analysis |  | Longitudinal Analysis |  |
|---|---|---|---|---|
|  | Friedreich's Ataxia | Control | Friedreich's Ataxia | Control |
| n Subjects | 57 | 38 | 35 | 25 |
| n Sessions (ankle) | 136 | 94 | 114 | 81 |
| n Sessions (wrist) | 136 | 95 | 114 | 82 |
| Female | 37 | 21 | 25 | 15 |
| Male | 20 | 17 | 10 | 10 |
| Median Age (years) | 28.8 (3.1–68.7) | 28.6 (4.8–70.5) | 31.0 (3.1–59.2) | 27.1 (4.8–70.5) |
| Median mFARS Total Score (N=22) | 42 (13–73) | NA | 42 (16–73) | NA |
| Median Duration of Participation (years) | --- | --- | 1.6 (0.5–2.2) | 1.6 (0.5–2.1) |
| Sessions Per Subject | --- | --- | 3 (2–5) | 3 (2–4) |
| Median GAA Repeats (n=46) | 700 (266–1183) | NA | 699 (266–990) | NA |
| Point Mutations | n=3 | NA | n=3 | NA |
| Skyclarys (omaveloxolone) | 37 (65%) | NA | 27 (77%) | NA |
| Median Duration of Symptoms (years) | 15.1 (0.2–37.2) | NA | 17.8 (1.6–37.2) | NA |
| Full Time Wheelchair | 27 (47%) | NA | 19 (54%) | NA |
| Part Time Wheelchair | 2 (3.5%) | NA | 1 (3%) | NA |
| Part Time Wheelchair + Another Assistive Device | 6 (10.5%) | NA | 4 (11%) | NA |
| Full Time Assistive Device (non-wheelchair) | 7 (12%) | NA | 4 (11%) | NA |
| Part Time Assistive Device (non-wheelchair) | 5(9%) | NA | 4 (11%) | NA |
| None | 10 (17.5%) | NA | 3 (9%) | NA |

Control participants did not complete mFARS. mFARS data are available only for FRDA participants enrolled in the UNIFAI natural history study at CHOP (N=22); all mFARS scores reflect baseline (T1) values and range from 0-93. GAA repeat lengths reflect Allele I (shorter allele). Duration of symptoms was calculated as age at enrollment minus age of symptom onset; one individual was presymptomatic and excluded from this calculation. Skyclarys (omaveloxolone) reflects the number of FRDA participants who were on the medication at any point during the study, out of those with Skyclarys status recorded. Assistive walking device data reflect the baseline timepoint for all participants. Walking device percentages are calculated among FRDA participants only. Ankle session data were used for sessions per subject and mean duration of participation calculations. FRDA: Friedreich’s ataxia; mFARS: modified Friedreich’s Ataxia Rating Scale; NA: not applicable; CHOP: Children’s Hospital of Philadelphia; UNIFAI: Friedreich Ataxia Global Clinical Consortium Unified Natural History Study; GAA: guanine, adenine, and adenine.

### Equipment and supplies

All participants received two GENEActiv wearable sensors to wear on the dominant wrist and ankle. Sensors recorded triaxial accelerometer data at 100 Hz continuously for one week.^9,21^

### Clinical Data

Clinical data used for comparison with remote wearable-derived measures were obtained through collaboration with the Children’s Hospital of Philadelphia (CHOP), a clinical site for the Friedreich’s Ataxia Global Clinical Consortium UNIFIED Natural History Study (UNIFAI).^27^

The clinical instruments collected included: FARS, mFARS, Friedreich’s Ataxia-Functional Disability Staging (FA-FDS), and Friedreich’s Ataxia-Activities of Daily Living (FA-ADL). The FA-ADL was completed by the participant or caregiver. All clinical data used for analysis were collected within six months of the corresponding wearables study timepoint.

### Questionnaires

FRDA study participants completed several patient-reported outcome measure (PROM) questionnaires at each time point, including Patient Global Impression of Change (PGIC), Friedreich’s Ataxia-Health Index (FA-HI), and Friedreich’s Ataxia Caregiver Reported-Health Index (FACR-HI).

The PGIC assessed the patient’s perception of change in disease severity since the last study timepoint. To align the time span reflected by PGIC with other data collected in this study, only timepoints 2 and greater were used in analysis. PGIC was analyzed at the response and subject level (see Supplementary Materials).

The FA-HI was administered to all participants aged 8 years or older at each timepoint. Only FA-HI data were used in analysis, as these data were available for the majority of participants (81), whereas FACR-HI data were only available for 9, precluding correlational analyses. Of the 86 participants eligible for the FA-HI based on age, 81 (94%) completed the survey.

### Wearable sensor data processing

Participants’ wearable sensor data were manually segmented into daytime and nighttime periods based on activity level visualized in the accelerometer recordings.^9,10,21,28^ Segmentation procedures are described in Supplementary Materials. Only daytime segments were included in the analysis. Continuous wrist and ankle sensor data were automatically processed and analyzed to derive 85 movement features, as previously described (see Supplementary Materials).^10,21,23^

Machine-learned composite models were applied to the extracted features from each week-long recording session. Five modelling strategies were separately applied to ankle and wrist sensor data (10 models total). Each model computed a weighted linear combination of the 85 movement features to generate an overall severity estimate. Three of the five models were trained on this FRDA dataset. One regression model was trained on individuals between the ages of 0 and 30 to predict FA-HI total score (“FRDA 0-30 model”, separately for ankle and wrist data). Another regression model was trained on individuals between 5 and 50 to predict FA-HI total score (FRDA 5-50 model). The third model was trained using the “pairwise model” approach,^23^ in which the model learned the steepest direction of disease change in feature space based on longitudinal data, without using clinician or patient-reported information (FRDA 5-50 pairwise model). These three modeling strategies were used to account for heterogeneity in age and disease severity across the FRDA population and to compare different approaches for creating composite scores. Although these models were trained on a subset of the population using cross validation, they were evaluated on the entire population.

The other two model types (2 ankle and 2 wrist) were previously trained on separate wearable sensor datasets from individuals with ALS^23^ (ALS pairwise model) and individuals with A-T (AT model). Additional model details are provided in the Supplementary Materials.

In addition to the composite models combining all 85 movement features, eight submovement (SM) summary features and one activity intensity (AI) feature (entropy of AI) were selected a priori for individual feature analysis and reporting. The nine features included the mean and standard deviation of SM duration, distance, peak velocity, and peak acceleration for short-duration SMs in the primary direction of movement. Statistical analyses for sensor-based measures are described in Supplementary Materials.

## Results

### Cross-sectional properties of ankle movement measures

Ankle sensor-derived composite models, submovement (SM) features, and activity intensity (AI) entropy were significantly correlated with clinician assessments including FA-FDS (*|r| =* 0.66 -0.87, Table 2), mFARS total (*|r| =* 0.53 -0.73), and mFARS upright stability (*|r| =* 0.58 -0.81). They also correlated significantly with self-reported FA-ADL (*|r| =* 0.58 -0.74), and FA-HI total (*|r| =* 0.43 -0.57), activity participation (*|r| =* 0.48 -0.66), and mobility & ambulation (*|r| =* 0.42 -0.57). AI entropy and the mean and standard deviation of SM distance, velocity, and acceleration all decreased with increasing disease severity, consistent with prior work in other ataxias.^9,10,21^ Mean SM duration (of short-duration SMs) increased with disease severity and showed the weakest correlations across all measures.

**Table 2.**
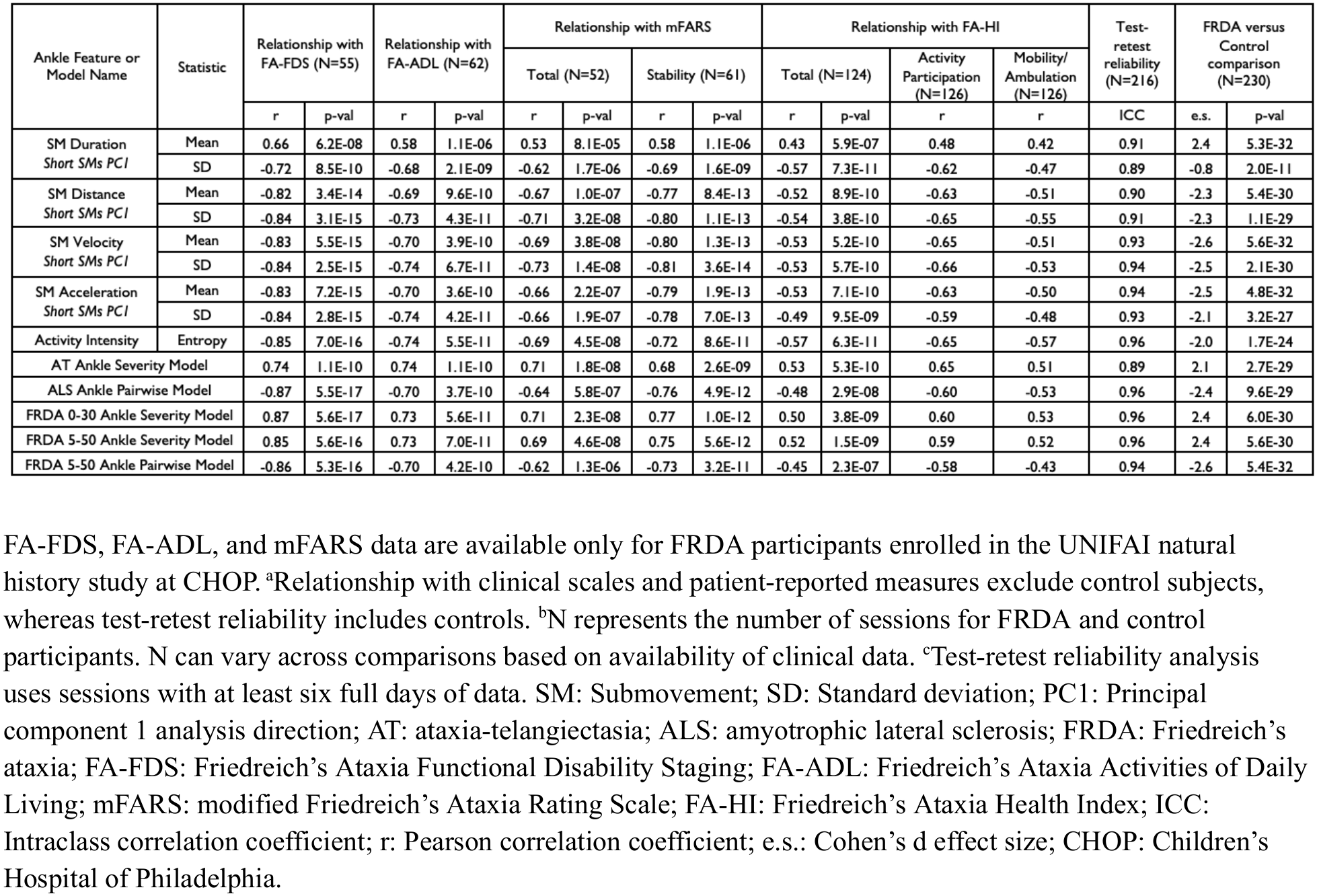
Relationship with scales and test-retest reliability of ankle sensor features and models.

Consistency analysis, comparing movement features obtained from the first versus second half of the week, demonstrated high reliability (*ICC* = 0.89 -0.96, Table 2). All movement features also differed strongly between FRDA and control populations (|*e.s.|* = 2.0 -2.6), except standard deviation of SM duration, which fell below an effect size of 1 (Table 2).

All five composite ankle models demonstrated similarly strong performance across cross-sectional evaluations, with the notable exception that the AT model had lower reliability (ICC = 0.89) in the FRDA population. Cross-sectional properties of the FRDA 0-30 severity model are shown in Figure 1.

**Figure 1.**
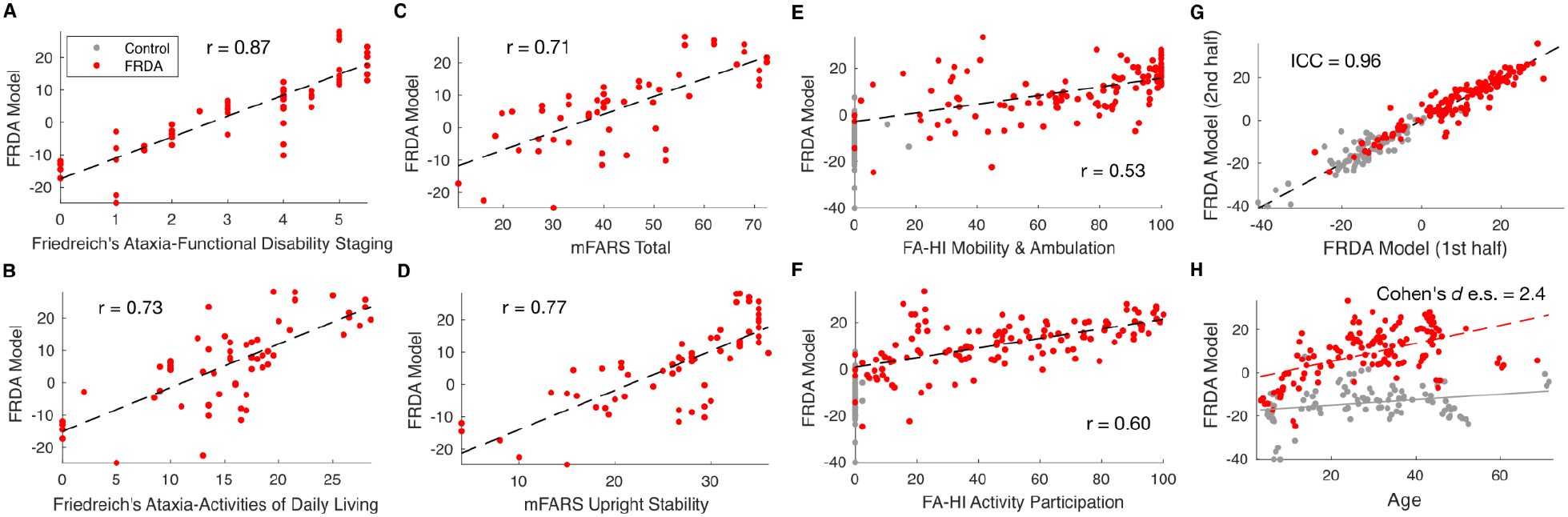
Cross-sectional properties of the ankle FRDA composite model. (A and B) Relationship of the FRDA composite model with FA-FDS and FA-ADL, respectively. (C and D) Relationship of the FRDA composite model to mFARS total and upright stability subscores, respectively. (E and F) Relationship of the FRDA composite model to FA-HI mobility and ambulation and activity participation, respectively. (G) Test-retest reliability of the FRDA composite model. (H) FRDA versus control scatter plot of the FRDA composite model. Each point represents one session. FRDA = Friedreich’s ataxia; e.s. = Cohen’s d effect size; ICC = Intraclass correlation coefficient; FA-HI = Friedreich’s Ataxia Health Index; FA-FDS = Friedreich’s Ataxia Functional Disability Staging; FA-ADL = Friedreich’s Ataxia Activities of Daily Living; mFARS = modified Friedreich’s Ataxia Rating Scale; r = Pearson correlation coefficient.

To understand which individual features were most salient in the FRDA 0-30 ankle severity model, we identified features that were in the top 10 (out of 85) in feature importance for at least 75% of the cross-validation folds. The top features included AI entropy, mean SM duration, and standard deviation of SM peak velocity (for short-duration SMs in the primary direction of motion). The model assigned higher severity scores to individuals with lower AI entropy and lower variance in SM peak velocity, and higher scores to individuals with longer SM durations. The model weights were consistent with individual feature analysis (Table 2) further supporting that natural everyday movement units become less variable and slower in individuals with _ataxia.9,10,21_

### Cross-sectional properties of wrist movement measures

Wrist sensor-derived submovement (SM) features, activity intensity (AI) entropy, and composite models were all significantly correlated with FA-FDS (*|r| =* 0.53 -0.83, Table 3), but only composite models correlated significantly with mFARS total (*|r| =* 0.43 -0.81) and mFARS upper limb (*|r| =* 0.35 -0.62), with the strongest performance observed for the FRDA 5-50 pairwise model (Table 3). Relationships with self-reported measures of function were significant but weaker on average than the ankle-derived features: FA-ADL (*|r| =* 0.30 -0.79), FA-HI total (*|r| =* 0.23 -0.49), FA-HI activity participation (*|r| =* 0.18 -0.55), and FA-HI shoulder and arm (*|r| =* 0.19 -0.35). Similar to the ankle-based measures, the mean and standard deviation of wrist SM distance, velocity, and acceleration all decreased with increasing disease severity.

**Table 3.** Relationship with scales and test-retest reliability of wrist sensor features and models.

| Wrist Feature or Model Name | Statistic | Relationship with FA-FDS (N=56) |  | Relationship with FA-ADL (N=63) |  | Relationship with mFARS |  |  |  | Relationship with FA-HI |  |  |  | Test-retest reliability (N=216) | FRDA versus Control comparison (N=231) |  |
|---|---|---|---|---|---|---|---|---|---|---|---|---|---|---|---|---|
|  |  |  |  |  |  | Total (N=53) |  | Arm (N=63) |  | Total (N=124) |  | Activity Participation (N=126) | Shoulder/Arm (N=126) |  |  |  |
|  |  | r | p-val | r | p-val | r | p-val | r | p-val | r | p-val | r | r | ICC | e.s. | p-val |
| SM Duration<br>*Short SMs PC1* | Mean | 0.53 | 3.5E-05 | 0.44 | 5.3E-04 | - | n.s. | - | n.s. | 0.42 | 4.4E-06 | 0.37 | 0.27 | 0.88 | 1.5 | 8.0E-19 |
|  | SD | -0.66 | 8.7E-08 | -0.48 | 1.6E-04 | - | n.s. | - | n.s. | -0.42 | 3.9E-06 | -0.39 | -0.25 | 0.84 | -1.0 | 3.8E-12 |
| SM Distance<br>*Short SMs PC1* | Mean | -0.49 | 1.3E-04 | -0.30 | 1.8E-02 | - | n.s. | - | n.s. | -0.23 | 9.8E-03 | -0.18 | - | 0.79 | -1.1 | 1.6E-14 |
|  | SD | -0.53 | 3.1E-05 | -0.33 | 8.8E-03 | - | n.s. | - | n.s. | -0.25 | 5.3E-03 | -0.23 | - | 0.64 | -1.0 | 1.1E-14 |
| SM Velocity<br>*Short SMs PC1* | Mean | -0.56 | 8.9E-06 | -0.39 | 2.0E-03 | - | n.s. | - | n.s. | -0.34 | 2.0E-04 | -0.27 | -0.22 | 0.84 | -1.2 | 1.1E-17 |
|  | SD | -0.60 | 2.0E-06 | -0.42 | 9.5E-04 | - | n.s. | - | n.s. | -0.30 | 9.9E-04 | -0.26 | -0.19 | 0.69 | -1.1 | 1.4E-16 |
| SM Acceleration<br>*Short SMs PC1* | Mean | -0.59 | 2.5E-06 | -0.42 | 8.5E-04 | - | n.s. | - | n.s. | -0.39 | 2.6E-05 | -0.31 | -0.27 | 0.84 | -1.0 | 2.6E-16 |
|  | SD | -0.50 | 1.1E-04 | -0.30 | 1.7E-02 | - | n.s. | - | n.s. | -0.26 | 4.5E-03 | -0.18 | -0.19 | 0.70 | -0.6 | 1.4E-06 |
| Activity Intensity | Entropy | -0.74 | 3.1E-10 | -0.55 | 1.2E-05 | -0.43 | 4.4E-03 | - | n.s. | -0.38 | 3.0E-05 | -0.45 | -0.27 | 0.95 | -1.5 | 1.0E-19 |
| AT Wrist Severity Model |  | 0.78 | 1.8E-11 | 0.67 | 1.7E-08 | 0.58 | 4.4E-05 | 0.35 | 2.5E-02 | 0.38 | 2.5E-05 | 0.53 | - | 0.93 | 2.2 | 1.2E-28 |
| ALS Wrist Pairwise Model |  | -0.77 | 2.0E-11 | -0.60 | 1.0E-06 | -0.49 | 8.9E-04 | - | n.s. | -0.49 | 2.0E-07 | -0.55 | -0.28 | 0.94 | -1.7 | 1.4E-21 |
| FRDA 0-30 Wrist Severity Model |  | 0.72 | 9.6E-10 | 0.54 | 1.9E-05 | 0.49 | 7.5E-04 | 0.36 | 3.6E-02 | 0.46 | 5.3E-07 | 0.50 | 0.35 | 0.85 | 2.2 | 1.6E-28 |
| FRDA 5-50 Wrist Severity Model |  | 0.74 | 2.3E-10 | 0.56 | 7.0E-06 | 0.53 | 2.8E-04 | 0.35 | 2.8E-02 | 0.46 | 6.3E-07 | 0.52 | 0.29 | 0.91 | 2.4 | 4.6E-29 |
| FRDA 5-50 Wrist Pairwise Model |  | -0.83 | 2.5E-14 | -0.79 | 1.6E-13 | -0.81 | 2.6E-12 | -0.62 | 1.5E-06 | -0.37 | 5.3E-05 | -0.50 | -0.28 | 0.88 | -2.8 | 8.1E-35 |

FA-FDS, FA-ADL, and mFARS data are available only for FRDA participants enrolled in the UNIFAI natural history study at CHOP. <sup>a</sup>Relationship with clinical scales and patient-reported measures exclude control subjects, whereas test-retest reliability includes controls. <sup>b</sup>N represents the number of sessions for FRDA and control participants. N can vary across comparisons based on availability of clinical data. <sup>c</sup>Test-retest reliability analysis uses sessions with at least six full days of data. SM: Submovement; SD: Standard deviation; PC1: Principal component 1 analysis direction; AT: ataxia-telangiectasia; ALS: amyotrophic lateral sclerosis; FRDA: Friedreich's ataxia; FA-FDS: Friedreich's Ataxia Functional Disability Staging; FA-ADL: Friedreich's Ataxia Activities of Daily Living; mFARS: modified Friedreich's Ataxia Rating Scale; FA-HI: Friedreich's Ataxia Health Index; ICC: Intraclass correlation coefficient; r: Pearson correlation coefficient; e.s.: Cohen's d effect size; CHOP: Children's Hospital of Philadelphia.

Consistency analysis demonstrated moderate to high reliability for most wrist features (*ICC* = 0.64 -0.95, Table 3) and all features differed significantly between FRDA and control populations (|*e.s.|* = 0.6 -2.8, Table 3).

### Longitudinal properties of ankle and wrist movement measures

Most ankle sensor-based composite models (4/5) and individual features (6/9) had statistically significant change over time in the FRDA population, with the mean to standard deviation ratio (|*MSDR*|) of participant slopes for each measure ranging from 0.28-0.89 (Table 4). The direction of change of all measures was consistent with disease progression, as it aligned with the valence of cross-sectional relationships with clinical severity and differences with the control population (e.g., FRDA participants moved further away from the control distribution over time). The distribution of slopes for most measures was significantly different between the FRDA and control populations, with moderate to strong effect sizes (|*e.s.|* = 0.56 -1.08) and none of the measures significantly changed over time in the control population (Table 4).

**Table 4.** Sensitivity to disease progression for ankle and wrist sensor features and models.

| Ankle Feature or Model Name | Statistic | Ankle Sensor |  |  |  |  |  | Wrist Sensor |  |  |  |  |  |
|---|---|---|---|---|---|---|---|---|---|---|---|---|---|
|  |  | FRDA (N=35) |  | Control (N=25) |  | FRDA vs Control Slopes |  | FRDA (N=35) |  | Control (N=25) |  | FRDA vs Control Slopes |  |
|  |  | MSDR | p-val | MSDR | p-val | e.s. | p-val | MSDR | p-val | MSDR | p-val | e.s. | p-val |
| SM Duration<br>*Short SMs PC1* | Mean | 0.52 | 2.0E-02 | - | n.s. | 0.67 | 1.9E-02 | - | n.s. | - | n.s. | - | n.s. |
|  | SD | - | n.s. | - | n.s. | - | n.s. | - | n.s. | - | n.s. | - | n.s. |
| SM Distance<br>*Short SMs PC1* | Mean | -0.43 | 1.5E-02 | - | n.s. | -0.75 | 4.6E-03 | - | n.s. | - | n.s. | - | n.s. |
|  | SD | -0.28 | 4.7E-02 | - | n.s. | -0.63 | 1.2E-02 | - | n.s. | - | n.s. | - | n.s. |
| SM Velocity<br>*Short SMs PC1* | Mean | -0.43 | 1.4E-02 | - | n.s. | -0.82 | 1.1E-03 | - | n.s. | - | n.s. | - | n.s. |
|  | SD | - | n.s. | - | n.s. | -0.74 | 9.1E-03 | - | n.s. | - | n.s. | - | n.s. |
| SM Acceleration<br>*Short SMs PC1* | Mean | -0.39 | 2.0E-02 | - | n.s. | -0.78 | 4.2E-03 | - | n.s. | - | n.s. | - | n.s. |
|  | SD | - | n.s. | - | n.s. | -0.77 | 6.4E-03 | - | n.s. | - | n.s. | - | n.s. |
| Activity Intensity | Entropy | -0.74 | 2.0E-03 | - | n.s. | -0.65 | 8.3E-03 | - | n.s. | - | n.s. | - | n.s. |
| AT Ankle/Wrist Severity Model |  | 0.45 | 2.2E-02 | - | n.s. | 0.56 | 1.5E-02 | - | n.s. | - | n.s. | - | n.s. |
| ALS Ankle/Wrist Pairwise Model |  | - | n.s. | - | n.s. | - | n.s. | - | n.s. | - | n.s. | - | n.s. |
| FRDA 0-30 Ankle/Wrist Severity Model |  | 0.79 | 1.8E-03 | - | n.s. | 1.08 | 1.3E-04 | - | n.s. | - | n.s. | - | n.s. |
| FRDA 5-50 Ankle/Wrist Severity Model |  | 0.67 | 1.5E-03 | - | n.s. | 1.08 | 8.0E-05 | - | n.s. | - | n.s. | - | n.s. |
| FRDA 5-50 Ankle/Wrist Pairwise Model |  | -0.89 | 8.0E-04 | - | n.s. | -0.65 | 5.3E-03 | -0.60 | 7.7E-03 | - | n.s. | - | n.s. |

<sup>a</sup>N represents the number of FRDA or control participants with longitudinal data. <sup>b</sup>Relationships that are not significant have effect sizes reported as “-“. <sup>c</sup>Relationships that are not significant are labeled as “n.s.”. SM: Submovement; SD: Standard deviation; PC1: Principal component 1 analysis direction; AT: ataxia-telangiectasia; ALS: amyotrophic lateral sclerosis; FRDA: Friedreich’s ataxia; MSDR: mean progression slope divided by the standard deviation of the progression slope across participants; e.s.: Cohen’s d effect size comparing FRDA and Control slope distributions.

The three FRDA-specific composite models had higher MSDR values and larger differences between FRDA and control slope distributions than the ALS and AT composite models (Table 4). Longitudinal properties of the FRDA 0-30 severity model are shown in Figure 2. Rate of change of the FRDA 0-30 severity model was not related to participant age (Figure 2C) or duration of participation in the study (Figure 2D).

**Figure 2.**
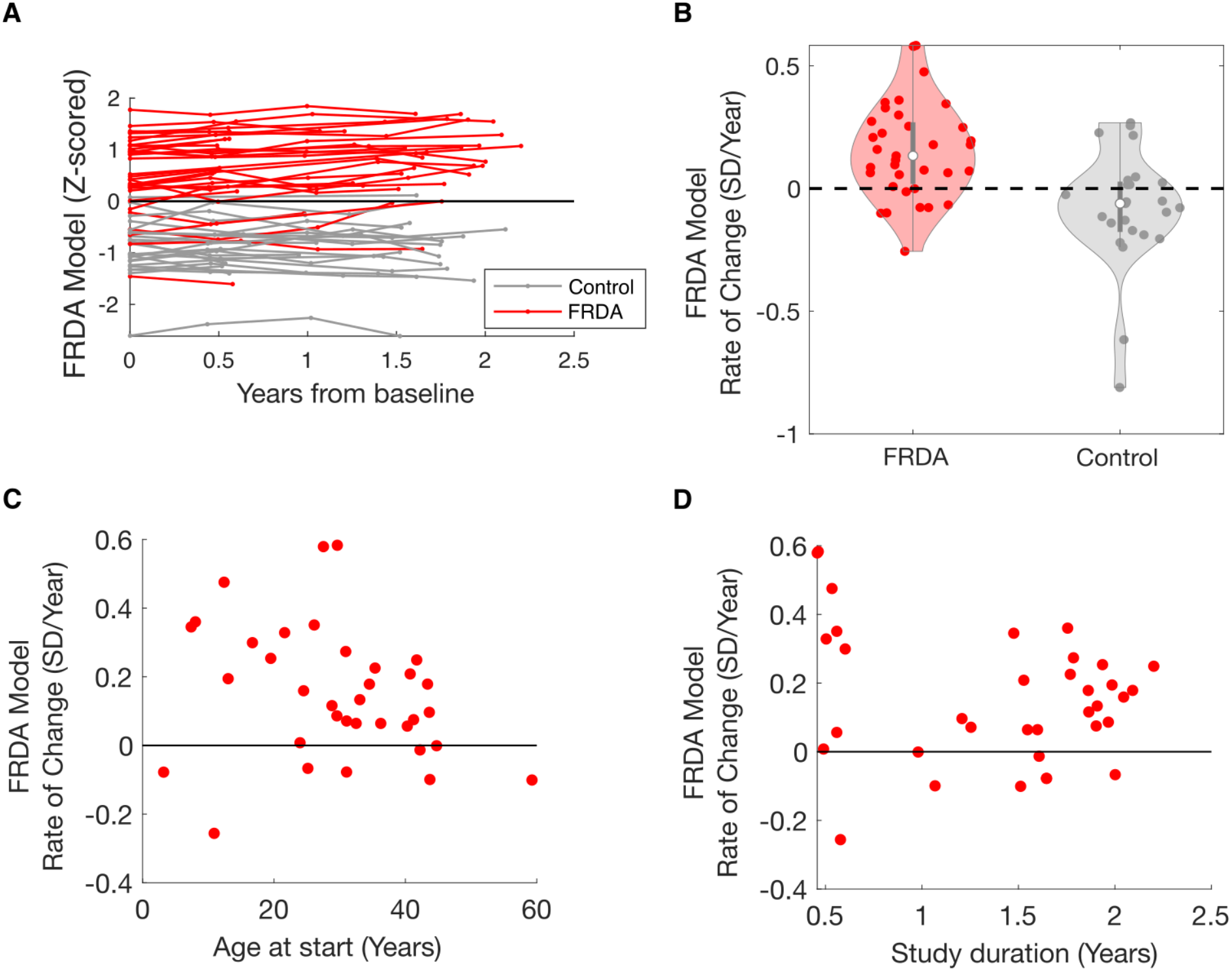
Longitudinal properties of the ankle FRDA composite model. (A) Change over time in the ankle FRDA composite model. Each line represents one participant, while each point represents one session. (B) Comparison of rates of change between FRDA and controls. The dashed line indicates zero rate of change. (C) FRDA model rate of change plotted against age at enrollment for FRDA participants. (D) FRDA model rate of change plotted against study duration for FRDA participants. Each point represents one participant. FRDA = Friedreich’s ataxia; SD = standard deviation.

For wrist sensor-based measures, only the FRDA 5-50 pairwise model demonstrated significant sensitivity to change in the FRDA population (|*MSDR*| = 0.60), however participant slopes were not significantly different between the FRDA and control populations (Table 4).

### Longitudinal properties of Patient Global Impression of Change (PGIC)

To compare the longitudinal properties of sensor-based measures with participant-reported change, we analyzed Patient Global Impression of Change (PGIC) data from the 35 individuals with FRDA who had longitudinal sensor data. A total of 82 responses were collected across timepoints 2-5 (T2 *n*=35, T3 *n*=26, T4 *n*=17, T5 *n*=4). Across these 82 responses, the majority indicated stability (*n*=47, 57%) followed by progression (*n*=20, 24%), and improvement (*n*=15, 18%). At the individual level, average PGIC scores across the 35 participants revealed a predominant trend of stability (average score=3; *n*=14), followed by progression (average score > 3, range 3.33-4.00; *n*=11), with slightly fewer participants reporting improvement (average score < 3, range 1.67-2.67; *n*=10). After subtracting 3 from each individual’s mean PGIC score (i.e., 0 indicates no change), the distribution of PGIC scores was not significantly different from a zero median distribution (p=0.94), indicating PGIC did not capture progression in the cohort.

## Discussion

We found that ecologically valid submovement-based measures, derived from accelerometer data collected at home, produce reliable and sensitive outcome measures in FRDA across a wide age and severity spectrum. Ankle sensor measures demonstrated strong cross-sectional correlations with clinician-rated and patient-reported outcomes, high within-week reliability, and large differences between FRDA and control groups. FRDA-specific machine-learned composite models showed the strongest sensitivity to longitudinal disease progression, which was not captured by PGIC, and outperformed models trained on data from other neurologic populations. Together, these findings support the potential of remote wearable monitoring as a sensitive digital outcome measure for FRDA clinical trials and for disease monitoring.

We observed strong cross-sectional correlations between ankle sensor measures and FA-FDS, with stepwise increases between each disease stage (Figure 1A), supporting that the ankle measures can capture motor function from early to late stages of disease in FRDA. Correlations with FA-HI further support the meaningfulness of the measures; however, the smaller effect sizes compared with functional staging may reflect the subjective nature of patient report.

Consistent with prior work in SCAs, ankle measures consistently outperformed wrist measures in cross-sectional and longitudinal analyses.^9,21^ Ankle-derived measures may be especially informative as lower limb dysfunction, including progressive gait impairment and loss of ambulation, is a defining feature of disease progression. However, approximately 50% of FRDA participants in this study (and in typical FRDA populations)^29^ are full time wheelchair users. This supports that the longitudinally sensitive ankle composite measures were able to capture lower limb impairment and progression beyond ambulation. By weighing multiple movement properties, including the range of activity intensity (AI entropy) and submovement characteristics that reflect movement quality, composite measures have the potential to accommodate a wider spectrum of disease than a single measure. Greater variability of upper limb activities during natural behavior may contribute to the weaker longitudinal performance of wrist measures. Consistent with natural history data, upper limb function declines at a slower rate than the lower limb in FRDA.^29,30^ Future work is needed to expand on the relative contributions of wrist and ankle sensor data for tracking FRDA progression in larger cohorts over a longer time. A larger longitudinal dataset would also enable improved training of machine-learned composite scores optimized for measuring disease change.^23^ The wrist pairwise model, trained to detect longitudinal change, was the only wrist measure that captured progression, underscoring the potential for improved performance with future modelling efforts.

The observed submovement properties in FRDA were consistent with prior work across other ataxia populations, including SCAs^9,21^ and A-T.^10,28^ Submovements in FRDA became smaller, slower, and less variable with increased severity, and AI entropy decreased with worsening motor impairment. These findings reflect the decomposition of movements into smaller, less coordinated segments,^9,14,17,21^ a hallmark of the ataxia phenotype, and a reduction in the range of movement intensities during daily activity. The consistency of submovement and activity intensity patterns across ataxias (FRDA, SCAs, A-T) and other neurological conditions (Parkinson disease^22^ and ALS)^23^, supports the generalizability of this analytic framework across populations with motor impairment.

All five composite models, including those trained in FRDA, A-T, and ALS datasets, demonstrated strong cross-sectional performance in FRDA, further supporting the overlap in ataxia phenotype, captured by wearable sensors, across these populations. However,

FRDA-specific models demonstrated stronger longitudinal sensitivity than models transferred from other populations, with higher MSDR values and larger differences between FRDA and control slope distributions. This difference indicates that while the cross-sectional movement features of ataxia may be broadly similar across diagnoses, the precise patterns of motor change and progression likely differ across ataxias. These results motivate additional longitudinal data collection in larger FRDA cohorts, including younger and earlier stage individuals, to further optimize FRDA-specific models. Notably, whereas the objective wearable sensor-based measures captured progression in FRDA, patient global impression of change (PGIC) did not, with the largest proportion of individuals reporting no change (*n*=14), and a nearly equal number reporting worsening (*n*=11) and improvement (*n*=10). This may reflect that individuals with FRDA experience change gradually and subtly over short intervals and/or that the immediate/short-term emotional, physical, and cognitive state of the individual (e.g., having a good day) contributes to their perception of disease change.

This study had several limitations. The cohort was relatively modest in size and spanned a wide range of ages and severity levels. The population included pediatric and adult participants, ambulatory and non-ambulatory individuals, and early through late-stage disease. While this breadth supports broad applicability, the modest sample size and heterogeneity limited detailed subgroup analysis. Clinical outcome data were available only for a subset of FRDA participants through collaboration with the UNIFAI natural history study, limiting the number of participants for whom wearable measures could be validated against clinician-administered assessments.

Future work may benefit from expanded longitudinal data collection, with a focus on younger and earlier stage individuals with FRDA who are underrepresented in the current cohort.

Submovement-based measures derived from continuous at-home accelerometer data, particularly from the ankle, produce reliable, ecologically valid, and sensitive outcome measures across a wide age and severity spectrum in FRDA. This is a promising approach to developing scalable digital outcome measures for FRDA clinical trials, and future work with larger and earlier-stage cohorts will enable further optimization of FRDA-specific models and broader applicability across the disease spectrum.

## Data Availability

De-identified participant data and a data dictionary will be made available to qualified investigators upon reasonable request to the corresponding author.

## Contributors

ASG contributed to the conception and design of the study. KA, FY, JF, MR, VP, and DL contributed to participant recruitment. KA and FY contributed to data acquisition and coordinated study visits. DL contributed clinical evaluations. MR and VP assisted clinical data transfers. KA and ASG assessed and verified the underlying data. ASG and KA performed the data analysis. ASG contributed to funding acquisition, supervision, and validation of the results. KA and ASG drafted the manuscript and prepared the figures. All authors critically reviewed the manuscript. All authors had full access to all the data in the study, reviewed and approved the final manuscript, and had final responsibility for the decision to submit for publication.

## Acknowledgements

We thank the participants and their families for their time and dedication to this research, and the Friedreich’s Ataxia Research Alliance (FARA) and investigators of the FA Global Clinical Consortium for their support with recruitment and participant referrals. We would also like to thank Dr. Chad Heatwole and his team at the University of Rochester for providing the FA-HI and FACR-HI surveys. This work was supported by FARA and the U.S. National Institutes of Health (R01 NS134597).

## AI Disclosure Statement

The authors used AI tools to suggest edits to condense the manuscript and improve clarity, identify grammatical errors, and identify additional literature of relevance.

## References

1. Scott V, Delatycki MB, Tai G, Corben LA. New and emerging drug and gene therapies for Friedreich Ataxia. CNS Drugs 2024;38(10):791–805.

2. Lynch DR, Chin MP, Delatycki MB, et al. Safety and efficacy of omaveloxolone in friedreich ataxia (MOXIe study). Ann. Neurol. 2021;89(2):212–225.

3. Reetz K, Lischewski SA, Dogan I, et al. Friedreich’s ataxia-a rare multisystem disease. Lancet Neurol. 2025;24(7):614–624.

4. Lynch DR, Farmer JM, Balcer LJ, Wilson RB. Friedreich ataxia: effects of genetic understanding on clinical evaluation and therapy. Arch. Neurol. 2002;59(5):743–747.

5. Pandolfo M. Friedreich ataxia. Arch. Neurol. 2008;65(10):1296–1303.

6. Subramony SH, Lynch DR. Friedreich ataxia. Pediatr. Neurol. 2026;174:148–154.

7. Brandsma R, Spits AH, Kuiper MJ, et al. Ataxia rating scales are age-dependent in healthy children. Dev. Med. Child Neurol. 2014;56(6):556–563.

8. Rummey C, Corben LA, Delatycki MB, et al. Psychometric properties of the Friedreich Ataxia Rating Scale. Neurol. Genet. 2019;5(6):371.

9. Manohar R, Yang FX, Stephen CD, et al. At-home wearables and machine learning capture motor impairment and progression in adult ataxias. Brain 2025;148(10):3623–3634.

10. Gupta AS, Luddy AC, Khan NC, et al. Real-life wrist movement patterns capture motor impairment in individuals with ataxia-telangiectasia. Cerebellum 2023;22(2):261–271.

11. Rummey C, Kichula E, Lynch DR. Clinical trial design for Friedreich ataxia -Where are we now and what do we need? Expert Opin. Orphan Drugs 2018;6(3):219–230.

12. Brooker SM, Edamakanti CR, Akasha SM, et al. Spinocerebellar ataxia clinical trials: opportunities and challenges. Ann. Clin. Transl. Neurol. 2021;8(7):1543–1556.

13. Mathiowetz V, Weber K, Kashman N, Volland G. Adult norms for the Nine Hole Peg Test of finger dexterity. Occup. Ther. J. Res. 1985;5(1):24–38.

14. Gupta AS. Digital phenotyping in clinical neurology. Semin. Neurol. 2022;42(1):48–59.

15. Stroud C, Onnela J-P, Manji H. Harnessing digital technology to predict, diagnose, monitor, and develop treatments for brain disorders. NPJ Digit. Med. 2019;2(1):44.

16. Hogan N, Sternad D. Dynamic primitives of motor behavior. Biol. Cybern. 2012;106(11–12):727–739.

17. Holmes G. The cerebellum of man. Brain 1939;62(1):1–30.

18. Oubre B, Daneault J-F, Whritenour K, et al. Decomposition of reaching movements enables detection and measurement of ataxia. Cerebellum 2021;20(6):811–822.

19. Lee J, Oubre B, Daneault J-F, et al. Analysis of gait sub-movements to estimate ataxia severity using ankle inertial data. IEEE Trans. Biomed. Eng. 2022;69(7):2314–2323.

20. Lee J, Oubre B, Daneault J-F, et al. Estimation of ataxia severity in children with ataxia-telangiectasia using ankle-worn sensors. J. Neurol. 2023;270(10):5097–5101.

21. Eklund NM, Ouillon J, Pandey V, et al. Real-life ankle submovements and computer mouse use reflect patient-reported function in adult ataxias. Brain Commun 2023;5(2):fcad064.

22. Gupta AS, Patel S. Wrist accelerometry and machine learning sensitively capture disease progression in prodromal Parkinson’s disease. NPJ Parkinsons Dis. 2025;11(1):171.

23. Gupta AS, Patel S, Premasiri A, Vieira F. At-home wearables and machine learning sensitively capture disease progression in amyotrophic lateral sclerosis. Nat. Commun. 2023;14(1):5080.

24. Milne SC, Kim SH, Murphy A, et al. The responsiveness of gait and balance outcomes to disease progression in Friedreich ataxia. Cerebellum 2022;21(6):963–975.

25. Fichera M, Nanetti L, Monelli A, et al. Accelerometer-based measures in Friedreich ataxia: a longitudinal study on real-life activity. Front. Pharmacol. 2024;15:1342965.

26. Mishra RK, Nunes AS, Enriquez A, et al. At-home wearable-based monitoring predicts clinical measures and biological biomarkers of disease severity in Friedreich’s Ataxia. Commun. Med. (Lond.) 2024;4(1):217.

27. UNIFAI: Friedreich Ataxia Global Clinical Consortium Unified Natural History Study. ClinicalTrials.gov Identifier [date unknown];

28. Khan NC, Pandey V, Gajos KZ, Gupta AS. Free-Living Motor Activity Monitoring in Ataxia-Telangiectasia. Cerebellum 2022;21(3):368–379.

29. Rummey C, Corben LA, Delatycki M, et al. Natural history of Friedreich ataxia: Heterogeneity of neurologic progression and consequences for clinical trial design. Neurology 2022;99(14):e1499–e1510.

30. Patel M, Isaacs CJ, Seyer L, et al. Progression of Friedreich ataxia: quantitative characterization over 5 years. Ann. Clin. Transl. Neurol. 2016;3(9):684–694.

