## Supplementary Materials for "Continuous at-home wearables and machine learning sensitively track function and progression in pediatric and adult Friedreich’s ataxia"

### Supplementary Information

#### Supplementary Tables and Figures

**Supplementary Table 1. Individual participant demographic and clinical information.**
Control participants did not complete mFARS or FA-HI. mFARS data are available only for FRDA participants enrolled in the UNIFAI natural history study at CHOP (N=22); all mFARS scores reflect baseline (T1) values and mFARS ranges from 0-93. FA-HI total scores were the minimum scores across completed timepoints. Assistive walking device data reflects baseline timepoint for all participants. Participants with two walking devices reported equal use of both devices at the baseline visit.

FRDA, Friedreich's ataxia; mFARS, Modified Friedreich's Ataxia Rating Scale; FA-HI, Friedreich's Ataxia Health Index; NA, not applicable; *n*: number; CHOP: Children's Hospital of Philadelphia.
*FRDA participants who were ineligible for FA-HI based on age criteria.

| Decade of Life | Sex | Diagnosis | *n* Timepoints | Span (years) | mFARS Total | FA-HI Total | Walking Device |
|---|---|---|---|---|---|---|---|
| 3rd | Male | FRDA | 4 | 2.05 | N/A | 16.5 | Wheelchair |
| 5th | Male | FRDA | 3 | 2.20 | 49.8 | 20.1 | Wheelchair |
| 3rd | Male | FRDA | 2 | 0.46 | 37.0 | 41.5 | Walker |
| 5th | Female | FRDA | 4 | 2.09 | 71.0 | 55.7 | Wheelchair |
| 4th | Male | FRDA | 4 | 1.77 | 18.3 | 17.4 | Cane/Walking Stick |
| 2nd | Female | FRDA | 5 | 1.98 | 40.0 | 0.0 | None |
| 4th | Female | FRDA | 4 | 1.86 | 72.5 | 72.2 | Wheelchair |
| 4th | Female | FRDA | 4 | 1.91 | 38.7 | 63.1 | Walker |
| 3rd | Female | FRDA | 4 | 1.97 | N/A | 52.7 | Wheelchair |
| 3rd | Female | Control | 1 | N/A | N/A | N/A | None |
| 3rd | Female | FRDA | 1 | N/A | N/A | 30.7 | Wheelchair |
| 3rd | Female | Control | 1 | N/A | N/A | N/A | None |
| 4th | Male | FRDA | 3 | 1.79 | N/A | 58.6 | Wheelchair |
| 3rd | Female | FRDA | 2 | 0.46 | N/A | 44.9 | Walker |
| 2nd | Male | FRDA | 1 | N/A | N/A | 0.0 | None |
| 5th | Female | Control | 4 | 1.79 | N/A | N/A | None |

| Decade of Life | Sex | Diagnosis | *n* Timepoints | Span (years) | mFARS Total | FA-HI Total | Walking Device |
|---|---|---|---|---|---|---|---|
| 2nd | Male | Control | 3 | 2.11 | N/A | N/A | None |
| 5th | Male | Control | 3 | 1.37 | N/A | N/A | None |
| 2nd | Male | Control | 4 | 1.75 | N/A | N/A | None |
| 2nd | Male | Control | 4 | 1.74 | N/A | N/A | None |
| 3rd | Female | Control | 2 | 1.75 | N/A | N/A | None |
| 1st | Female | FRDA | 3 | 1.75 | 52.3 | 20.1 | Wheelchair |
| 6th | Female | FRDA | 4 | 1.51 | N/A | 51.4 | Walker & Wheelchair |
| 5th | Female | Control | 4 | 1.88 | N/A | N/A | None |
| 5th | Female | FRDA | 4 | 1.90 | N/A | 54.0 | Wheelchair |
| 2nd | Female | FRDA | 1 | N/A | 33.0 | 27.0 | Cane/Walking Stick |
| 2nd | Male | FRDA | 4 | 1.94 | N/A | 33.5 | Walker & Wheelchair |
| 3rd | Male | Control | 2 | 0.56 | N/A | N/A | None |
| 2nd | Female | FRDA | 2 | 0.60 | N/A | 16.5 | None |
| 5th | Female | FRDA | 1 | N/A | N/A | 58.8 | Wheelchair |
| 3rd | Female | FRDA | 3 | 1.86 | 47.0 | 44.0 | Wheelchair |
| 4th | Male | Control | 4 | 1.75 | N/A | N/A | None |
| 6th | Female | Control | 4 | 1.94 | N/A | N/A | None |
| 4th | Female | FRDA | 4 | 1.65 | N/A | 69.2 | Wheelchair |
| 3rd | Female | FRDA | 1 | N/A | N/A | 29.2 | Wheelchair |
| 5th | Female | Control | 1 | N/A | N/A | N/A | None |
| 2nd | Male | Control | 1 | N/A | N/A | N/A | None |
| 5th | Male | Control | 1 | N/A | N/A | N/A | None |
| 3rd | Male | FRDA | 2 | 0.49 | 55.0 | 29.2 | Wheelchair |
| 3rd | Female | Control | 2 | 0.67 | N/A | N/A | None |

| Decade of Life | Sex | Diagnosis | *n* Timepoints | Span (years) | mFARS Total | FA-HI Total | Walking Device |
|---|---|---|---|---|---|---|---|
| 3rd | Male | Control | 4 | 1.61 | N/A | N/A | None |
| 5th | Female | Control | 1 | N/A | N/A | N/A | None |
| 5th | Male | Control | 1 | N/A | N/A | N/A | None |
| 3rd | Female | FRDA | 2 | 0.50 | 33.0 | 39.6 | None |
| 2nd | Male | Control | 1 | N/A | N/A | N/A | None |
| 3rd | Female | Control | 2 | 1.61 | N/A | N/A | None |
| 1st | Female | Control | 3 | 1.61 | N/A | N/A | None |
| 3rd | Male | Control | 1 | N/A | N/A | N/A | None |
| 3rd | Female | Control | 4 | 1.57 | N/A | N/A | None |
| 2nd | Male | FRDA | 1 | N/A | N/A | 30.7 | Walker |
| 5th | Female | FRDA | 4 | 1.61 | 62.0 | 68.4 | Wheelchair |
| 4th | Female | FRDA | 4 | 1.60 | N/A | 31.4 | Walker & Wheelchair |
| 3rd | Male | FRDA | 5 | 2.00 | N/A | 74.3 | Wheelchair |
| 4th | Female | FRDA | 3 | 1.55 | N/A | 16.0 | Wheelchair |
| 1st | Female | FRDA | 4 | 1.48 | 44.5 | 19.8 | Orthosis & Walker |
| 1st | Male | FRDA | 4 | 1.65 | N/A | * | None |
| 1st | Female | Control | 4 | 1.70 | N/A | N/A | None |
| 4th | Female | Control | 4 | 1.53 | N/A | N/A | None |
| 5th | Male | FRDA | 1 | N/A | 57.0 | 50.5 | Wheelchair |
| 8th | Female | Control | 3 | 0.99 | N/A | N/A | None |
| 1st | Female | Control | 3 | 0.99 | N/A | N/A | None |
| 3rd | Female | FRDA | 2 | 0.56 | N/A | 42.6 | Wheelchair |
| 1st | Male | Control | 4 | 1.52 | N/A | N/A | None |
| 1st | Female | Control | 4 | 1.52 | N/A | N/A | None |
| 5th | Female | FRDA | 2 | 1.21 | N/A | 35.7 | Wheelchair |

| Decade of Life | Sex | Diagnosis | *n* Timepoints | Span (years) | mFARS Total | FA-HI Total | Walking Device |
|---|---|---|---|---|---|---|---|
| 2nd | Female | FRDA | 2 | 0.53 | N/A | 61.7 | Wheelchair |
| 5th | Female | FRDA | 4 | 1.53 | 31.3 | 34.6 | Cane/Walking Stick |
| 4th | Female | FRDA | 3 | 1.25 | N/A | 57.4 | Walker & Wheelchair |
| 3rd | Female | FRDA | 1 | N/A | N/A | 28.8 | None |
| 5th | Female | FRDA | 3 | 0.98 | 27.0 | 39.1 | Cane/Walking Stick |
| 4th | Male | Control | 3 | 0.95 | N/A | N/A | None |
| 5th | Female | FRDA | 3 | 1.07 | N/A | 31.3 | Wheelchair |
| 4th | Male | Control | 2 | 0.52 | N/A | N/A | None |
| 3rd | Female | Control | 3 | 1.05 | N/A | N/A | None |
| 4th | Female | Control | 1 | N/A | N/A | N/A | None |
| 4th | Male | Control | 1 | N/A | N/A | N/A | None |
| 2nd | Male | FRDA | 1 | N/A | 27.0 | 42.1 | None |
| 4th | Female | Control | 2 | 0.54 | N/A | N/A | None |
| 5th | Female | FRDA | 2 | 0.56 | N/A | 63.3 | Wheelchair |
| 1st | Male | FRDA | 1 | N/A | N/A | 11.9 | None |
| 1st | Male | FRDA | 1 | N/A | 13.0 | * | None |
| 2nd | Male | FRDA | 2 | 0.58 | 16.0 | 6.6 | Walker |
| 2nd | Male | FRDA | 1 | N/A | N/A | 44.9 | Wheelchair |
| 1st | Female | FRDA | 1 | N/A | N/A | * | None |
| 6th | Female | FRDA | 1 | N/A | N/A | 45.8 | Wheelchair |
| 3rd | Male | FRDA | 1 | N/A | N/A | 26.1 | Walker |
| 2nd | Female | FRDA | 1 | N/A | 51.0 | 40.7 | Wheelchair |
| 7th | Female | FRDA | 1 | N/A | N/A | 51.8 | Walker & Wheelchair |

| Decade of Life | Sex | Diagnosis | *n* Timepoints | Span (years) | mFARS Total | FA-HI Total | Walking Device |
|---|---|---|---|---|---|---|---|
| 4th | Female | Control | 1 | N/A | N/A | N/A | None |
| 3rd | Female | FRDA | 1 | N/A | N/A | 42.7 | Wheelchair |
| 3rd | Male | Control | 1 | N/A | N/A | N/A | None |
| 2nd | Male | FRDA | 1 | N/A | 44.0 | 45.3 | Walker & Wheelchair |
| 4th | Male | FRDA | 1 | N/A | N/A | 74.0 | Wheelchair |
| 4th | Female | FRDA | 1 | N/A | N/A | 42.6 | Wheelchair |
| 4th | Female | FRDA | 1 | N/A | N/A | 66.5 | Walker |

**Supplementary Figure 1. Flow chart describing the subset of total participants included in analysis.**

Of the 103 enrolled participants, 60 completed at least two timepoints and were included in longitudinal analysis. Of the 35 participants without longitudinal data, 13 (11 FRDA, 2 controls) had not yet reached their follow-up date by the end of data collection in January 2026. Of the remaining 22, 6 withdrew from the study: 4 controls cited scheduling constraints, 1 pediatric FRDA participant did not enjoy wearing the sensors, and 1 control withdrew due to a major life event. The remaining 16 participants (10 FRDA, 6 controls) were lost to follow-up after two attempted contacts by the study coordinator at consecutive target dates. FRDA, Friedreich's ataxia

*Divorce

[a]Unable to complete study tasks due to limited limb movement and visual impairment.

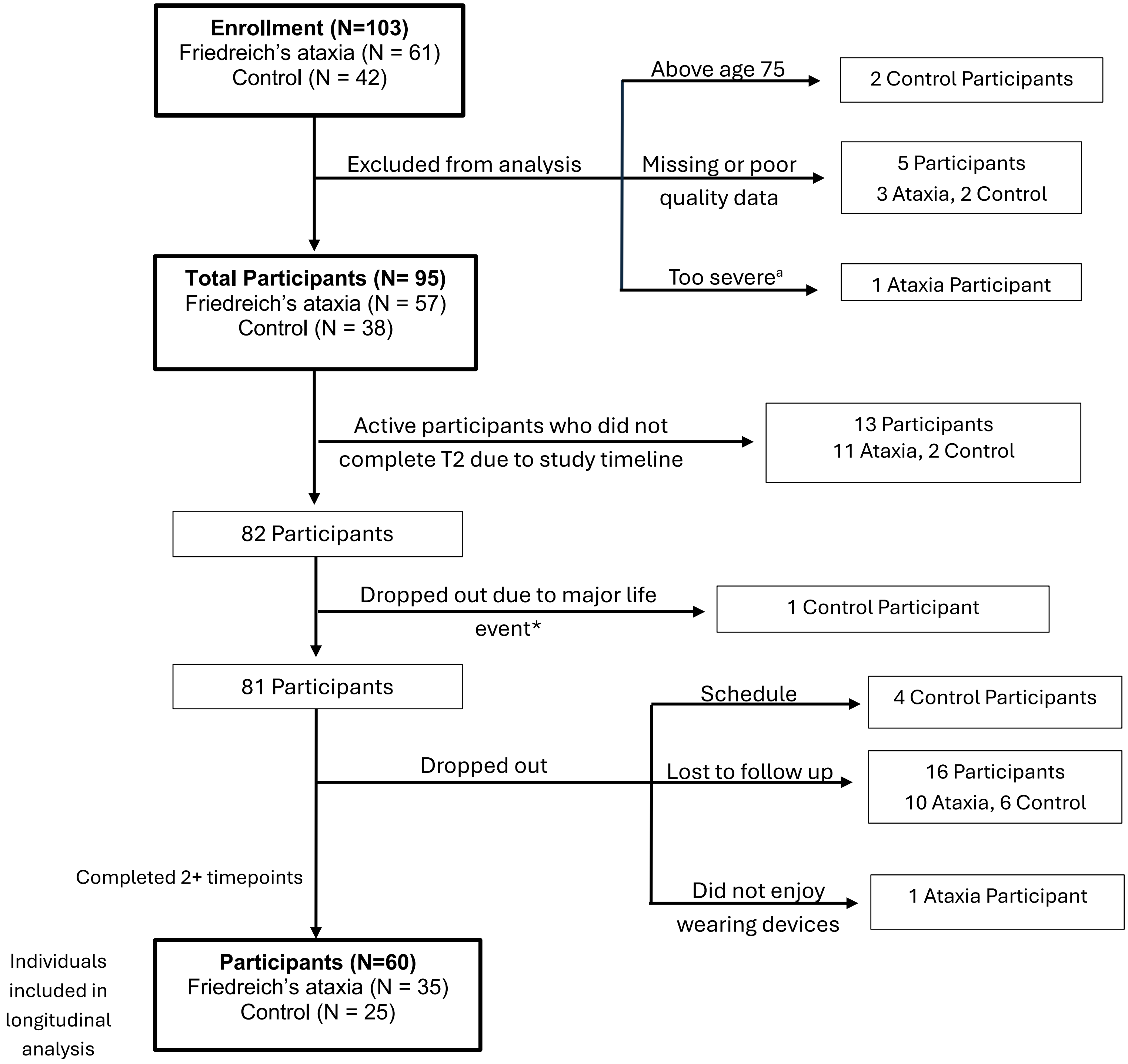

#### Supplementary Methods

#### Recruitment and Consent

Study participants were recruited mainly from the Friedreich's Ataxia Research Alliance (FARA) websites, social media, and patient registries. Healthy controls were recruited primarily through participant referrals, including siblings, spouses, other relatives, and friends. Controls were screened for neurological, musculoskeletal, or other conditions that may affect motor or cognitive function.

All 57 Friedreich's ataxia participants had prior genetic testing confirming their diagnosis. Of these, 48 provided their detailed genetic mutation information, while 9 did not have access to their results or were unable to provide the specific details of their mutation. Of the 48 who provided their results, 2 only knew a range of their Allele 1 GAA expansion repeat length, where Allele 1 is defined as the shorter of the two GAA repeat alleles; and 2 participants had a point mutation. One participant had a point mutation but did not know the details of their mutation.

#### Clinical Data

In addition to FARS[1] and mFARS[2] scores, clinical data for FRDA participants was obtained in collaboration with CHOP via the Friedreich's Ataxia-Functional Disability Staging (FA-FDS)[1] and Friedreich's Ataxia-Activities of Daily Living (FA-ADL)[1,2] instruments. The Timed 25-Foot Walk (T25-FW), an assessment used alongside FARS/mFARS,[1,2] is referenced in the main text but was not analyzed in this study.

#### Questionnaires

Questionnaires completed by FRDA participants at each study timepoint included the Pittsburgh Sleep Quality Index,[3] Children's Sleep Habits Questionnaire,[4] Patient Global Impression of Change (PGIC),[5] Friedreich's Ataxia-Health Index (FA-HI),[6] and if applicable, Friedreich's Ataxia Caregiver Reported-Health Index (FACR-HI).[7]

Of the 35 individuals with FRDA with two or more timepoints of wearable sensor data collection, all 35 were included in PGIC analysis. 34 had PGIC data across all wearable sensor timepoints (timepoint 2 and greater); one participant with 5 timepoints was missing a PGIC response for time point 4 and was included using their available responses. For analysis of PGIC, data were evaluated at the response and subject level. At the response level, PGIC responses from longitudinal timepoints (2-5) were compiled across all participants and the proportions of responses indicating improvement, stability, or progression were calculated. At the subject level, an average

PGIC score was calculated per participant using longitudinal PGIC responses, and these scores were used to assign participants to a corresponding trend: improvement (average scores below 3), stability (average score of 3), or progression (average scores above 3).

Prior to July 2025, the FACR-HI was administered only to participants between ages 5 and 8. Beginning in July 2025, the FACR-HI was expanded to include participants aged 8 and older who reported having a caregiver. Age thresholds for survey administration were determined based on guidance from the University of Rochester.

Five FA-HI entries were missing: four were due to participants not completing the survey, and one was due to a technical error in REDCap that prevented the survey invitation from being sent. All 9 participants eligible for the FACR-HI based on age completed the survey.

#### Wearable sensor data processing

Day/night segmentation for participants' wearable sensor data was initiated at sleep onset during the first complete night of recording. This produced a maximum of six consecutive 24-hour periods of recording.

An automated pipeline processed continuous triaxial accelerometer data to identify 'activity bouts', defined as brief periods of uninterrupted movement lasting 4-18 seconds.[8] Next, the acceleration time series was converted to a velocity time series by integration. The triaxial velocity time series was subsequently projected onto a two-dimensional plane using principal component analysis to determine the primary and secondary directions of motion during each bout.[8,9,10] Submovements (i.e., typically bell-shaped velocity-time curves flanked by zero velocity crossings) were identified in the primary and secondary directions of movement and were grouped into short and long-duration submovements (SM). In total, 85 features were generated at different stages of the pipeline. These included total power in the 0.1-5 Hz frequency range (one feature), features based on the distribution of 'activity intensity' (AI) computed from one-second time bins (10 features), 'activity bout' kinematic properties (four features), SM shape properties (30 features), and SM kinematic properties (40 features).[8,9,10] The five SM kinematic properties were duration, distance, peak velocity, peak acceleration, and jerk. For each of the five SM kinematic properties, mean and standard deviation (two statistics) were computed for short-duration and long-duration SMs in the primary and secondary direction of movement (four SM groups) yielding eight features per property and 40 SM kinematic features overall.

For model training, linear regression models with L1 regularization (i.e. lasso regression)[11] were used in order to select a small subset of the input variables.[9] Each feature was *Z*-score transformed prior to model training such that feature value ranges and model weights were comparable. FA-HI total score was used as the target variable for the FRDA 0-30 and FRDA 5-50 models because it was available for the majority of individuals. Leave-one-subject-out cross-validation was used to train and estimate generalization performance of these models.

The ALS models were trained using the pairwise model approach described above and were utilized in this study as they were trained on the largest longitudinal wearable sensor dataset available in any population and demonstrated strong cross-sectional and longitudinal performance in ALS[10] as well as in adult ataxias.[12] The AT models were regression models trained to predict the Brief Ataxia Rating Scale total score based on data from individuals with AT.[8,13] The AT models were used in this study as A-T is the second most common autosomal recessive cerebellar ataxia (after FRDA) and has some overlap with FRDA in age range and phenotype. Although the models trained on FRDA data were evaluated using cross-validation and approximate generalization performance, models based on data collected from entirely separate studies in other disease populations, at times using different wearable devices collecting data at different sampling rates, enables a more comprehensive assessment for how the digital assessment technology may be able to generalize to new studies and contexts.

In addition to the composite models, nine features were selected a priori for individual feature analysis and reporting as described in the main text. These features were selected in order to survey each SM kinematic property and an AI-based feature which were previously shown to capture relevant information in other ataxia populations.[12,8,9] These features were included in analysis to provide a small set of interpretable single features for comparison with the composite models, which were trained to combine information from the full set of 85 movement features.

#### Statistical Analysis

Statistical analyses were completed in MATLAB version R2022a (Mathworks, Natick, MA). The Mann-Whitney *U*-test was used to determine sensor measure differences between FRDA and control groups, and Cohen's *d* was used to measure effect size. Signed effect sizes were reported, with negative values indicating that the FRDA group mean was smaller than the control group mean. The Benjamini-Hochberg method was used to adjust for multiple comparisons and corrected p-values were reported.[14] Corrected p-values < 0.05 were considered significant. To evaluate reliability of wrist and ankle features, features were computed from data recorded on days 1-3 and days 4-6, separately, and single-measure intraclass correlation coefficients (ICCs) were computed using a two-way mixed effects model.[15] Pearson correlation coefficients and p-values were used to evaluate the relationship between ankle and wrist sensor features with patient-reported measures of function (FA-HI total score and subscores) and clinician performed assessments including mFARS total and subscores, FA-ADL, and FA-FDS. As above, the Benjamini-Hochberg method was used to adjust for multiple comparisons for each sensor type.[14] To avoid inflated correlation values driven by differences between control and FRDA participants, Pearson correlation coefficients were computed using data from ataxia participants only. For longitudinal data analysis, each participant's progression rate for a given outcome was determined by fitting a linear regression model to the individual's longitudinal data for the outcome and using the slope of the curve to represent progression over time.[16] The mean and standard deviation of the slope for each outcome were computed across the population, and the mean to standard deviation ratio (MSDR) was used to report effect size for sensitivity to change. The Wilcoxon signed-rank test was used to determine if the outcome's slope was significantly different from zero and the Mann-Whitney *U*-test was used to test if FRDA slopes differed from control slopes. For longitudinal analysis of PGIC, which directly queries change between two consecutive time points, the average PGIC response was computed for each individual with longitudinal data to represent the mean rate of change. As a value of 3 represents no change, 3 was subtracted from each individual's rate of change such that a value of 0 represents no change. The signed-rank test was then used to determine if mean PGIC responses across the longitudinal cohort were different from zero. Corrected p-values $< 0.05$ were considered significant.

#### References


1. Subramony SH, May W, Lynch D, et al. Measuring Friedreich ataxia: Interrater reliability of a neurologic rating scale. Neurology 2005;64(7):1261–1262.

2. Lynch DR, Farmer JM, Tsou AY, et al. Measuring Friedreich ataxia: complementary features of examination and performance measures. Neurology 2006;66(11):1711–1716.

3. Buysse DJ, Reynolds CF 3rd, Monk TH, et al. The Pittsburgh Sleep Quality Index: a new instrument for psychiatric practice and research. Psychiatry Res. 1989;28(2):193–213.

4. Owens JA, Spirito A, McGuinn M. The Children's Sleep Habits Questionnaire (CSHQ): psychometric properties of a survey instrument for school-aged children. Sleep 2000;23(8):1043–1051.

5. Farrar JT, Young JP Jr, LaMoreaux L, et al. Clinical importance of changes in chronic pain intensity measured on an 11-point numerical pain rating scale. Pain 2001;94(2):149–158.

6. Seabury J, Rosero S, Varma A, et al. Friedreich's Ataxia-Health Index: Development and validation of a novel disease-specific patient-reported outcome measure. Neurol. Clin. Pract. 2023;13(5):e200180.

7. Seabury J, Varma A, Weinstein J, et al. Friedreich Ataxia Caregiver-Reported Health Index: Development of a novel, disease-specific caregiver-reported outcome measure. Neurol. Clin. Pract. 2024;14(3):e200303.

8. Gupta AS, Luddy AC, Khan NC, et al. Real-life Wrist Movement Patterns Capture Motor Impairment in Individuals with Ataxia-Telangiectasia. Cerebellum 2023;22(2):261–271.

9. Eklund NM, Ouillon J, Pandey V, et al. Real-life ankle submovements and computer mouse use reflect patient-reported function in adult ataxias. Brain Commun 2023;5(2):fcad064.

10. Gupta AS, Patel S, Premasiri A, Vieira F. At-home wearables and machine learning sensitively capture disease progression in amyotrophic lateral sclerosis. Nat. Commun. 2023;14(1):5080.

11. Tibshirani R. Regression shrinkage and selection via the lasso. J. R. Stat. Soc. 1996;58(1):267–288.

12. Manohar R, Yang FX, Stephen CD, et al. At-home wearables and machine learning capture motor impairment and progression in adult ataxias. Brain 2025;148(10):3623–3634.

13. Khan NC, Pandey V, Gajos KZ, Gupta AS. Free-Living Motor Activity Monitoring in Ataxia-Telangiectasia. Cerebellum 2022;21(3):368–379.

14. Benjamini Y, Hochberg Y. Controlling the false discovery rate: A practical and powerful approach to multiple testing. J. R. Stat. Soc. 1995;57(1):289–300.

15. Shrout PE, Fleiss JL. Intraclass correlations: Uses in assessing rater reliability. Psychol. Bull. 1979;86(2):420–428.

16. Shefner JM, Liu D, Leitner ML, et al. Quantitative strength testing in ALS clinical trials. Neurology 2016;87(6):617–624.[cited 2024 Aug 22 ]
